# Unpacking COVID-19 and Conspiracy Theories in the UK Black Community

**DOI:** 10.1101/2022.02.12.22270438

**Authors:** Tushna Vandrevala, Jane Hendy, Kristin Hanson, Lailah Alidu, Aftab Ala

## Abstract

**Objectives:** Conspiracy theories are associated with significant COVID-19 health consequences including lower engagement with protective behaviours. This study uses sensemaking theory, a process of constructing meanings through interpersonal exchanges that enable people to interpret their world to explain the theoretical process underlying the development of conspiratorial beliefs around COVID-19 within Black African and Caribbean communities in the UK.

**Design:** Qualitative, in-depth interviews were used.

**Methods:** Twenty-eight members of the communities were recruited; semi-structured interviews were analysed using grounded theory.

**Results:** Our findings provide an explanation of how an environment of crisis combined with current and historical mistrust, perceived injustice and inequality provided a context in which alternative conspiracy narratives could thrive. The nature of these conspiratorial beliefs made more sense to many of our respondent’s than institutional sources (such as the UK Government). Critically, these alternative beliefs helped respondents shape their decision-making, leading to non-engagement with COVID protective behaviours.

**Conclusions:** We conclude that the uncertainty of the pandemic, combined with historical and contemporary perceived injustice and mistrust, and a lack of specific identity-aligned messaging, created a perfect environment for conspiratorial sense-making to thrive. This alternative sensemaking was inconsistent with the health-protection messaging espoused by Government. To ensure all groups in society are protected, and for health promotion messages to take purchase, the experiences of different target audiences must be taken into account, with sensemaking anchored in lived experience.

## Introduction

Conspiratorial beliefs related to COVID-19, have been found to be more prevalent in ethnic and migrant communities (EMGs) and are associated with significant COVID-19 health consequences through lower levels of engagement with protective behaviours; including mask wearing, testing, and vaccine intention (Allington et al., 2021; Oleksy et al., 2021; Romer & Jamieson, 2020). Lower levels of engagement with COVID-19 testing are consistent with studies related to EMGs for other preventative health measures, such as HIV (Harb et al., 2019) and HPV (Harrington et al., 2021). The current study seeks to understand why the UK Black community (Black African and Black Caribbean communities) are less inclined to take up protective health behaviours. More specifically, we aim to develop a theory explaining the relationship between the development of conspiratorial beliefs around COVID-19 and such behaviours. Understanding this relationship and the development and nature of conspiracy beliefs is important for practitioners who are striving to communicate and create interventions to promote COVID health protective behaviours; such as vaccination up-take and testing, which has been lower in Britain’s Black community compared to other minority ethnic groups (Office for National Statistics, 2021; Vandrevala et al., 2022).

Research that seeks to explain health behaviours, including studies of conspiracy belief development have focused on majority populations, so do not specifically account for the differing lived experiences of minority groups (Butter & Knight, 2015; Druckman et al, 2021). In this study we capture the lived experience and the broader social context in which respondents live by employing a qualitative, process-based paradigm. We also utilise a theory that synthesises sociological and social psychological theories, to re-position understandings (our sensemaking) as an ongoing process, highly situated in lived experience and cultural and political contexts (Maitlis & Sonenshein, 2010; Weick, 1995).

Our findings provide an explanation of how an environment of crisis combined with current and historical mistrust, perceived injustice and inequality provided a context in which alternative conspiracy narratives could thrive. The nature of these conspiratorial beliefs made more sense to many of our respondent’s than institutional sources (such as the UK Government). Critically, these alternative beliefs helped respondents shape their decision-making, leading to non-engagement with COVID protective behaviours.

### Sensemaking and conspiratorial beliefs

For EMGs, the uncertainty caused by the pandemic was keenly felt. The news that ethnic minority groups in the UK were disproportionately affected by the COVID-19 virus came early (e.g., Barr et al., 2020), precipitating not only a health threat, but also a group identity threat by casting EMGs as more vulerable to COVID-19 (Hanson et al., 2021; Vandrevala et al., 2022), with the uncertainty of the pandemic was exacerbated by factors such as increased exposure to the virus because of front-line jobs, and more densely populated living quarters.

In such uncertain and threatening times people seek to reduce feelings of anxiety through various sources of meaning including information-seeking and narrative construction (van Prooijen & Douglas, 2017). Sensemaking is a process of this social construction of meaning, and is enacted when discrepant information or a ‘gap’ in understanding of the world interrupts normal life. The presence of the pandemic created one of these ‘gaps’ and was felt throughout the general population. To resolve the cognitive disorder created from these sensemaking gaps or ‘fault lines’ (Balogun & Johnson, 2005) the development of new meaning (that rationalises what is going on) was urgently needed (Weick, 1995). As an unexpected and threatening event, the pandemic is likely to have induced feelings of personal uncertainty and loss of control in much of the population (Whitson et al., 2015).

Central to this development of new meaning are external cues from the world around. Often these external cues are taken from stories and messages put out by others. Such sensemaking cures are rarely static, single authored or scripted; they constantly change with the moving external landscape (Näslund & Pemer, 2012). The vagaries of this process and the changing context can also lead to confusion with a lack of consensus emerging, regarding the best way forward (Gioia et al., 1994), creating inertia or alternatively allowing competing information or narratives to gain purchase.

In such an environment conspiracy theory can prosper (van Bavel et al., 2020). Conspiracy theories are beliefs that significant events are the result of malevolent actions from powerful groups who ‘pull the strings’ (Douglas et al., 2017). Such beliefs tend to be stronger during times of uncertainty, such as a pandemic. Although theories surrounding the existence, causes, and treatments related to the virus existed in the wider population (Miller, 2020; van Bavel et al., 2020), African-Americans tended to hold significantly more misperceptions, and UK EMGs twice as likely as those in identifying as White to believe in COVID-19-related conspiracy theories’ (Allington et al., 2021).

The extent to which these new messages or cues ‘hit home’ to form the foundation of new sensemaking is mediated by a person’s existing frames of reference, the context or foundation of their current life. Any new information needs to link or overlap in some way to existing courses of action, ways of thinking and interactions. So, the content of new information is mediated by existing knowledge, creating scope for intended and unintended consequences (Balogun & Johnson, 2005). The sensemaking paradigm therefore posits that the extent to which EMGs would find COVID-related messages plausible would be determined by the extent to which they see the messages, as coherent with their worldviews and lived experiences.

This is in contrast to health behaviour models such as the theory of planned behaviour (Ajzen, 1991) and the health belief model (Rosenstock, 1966) where health behaviour is conceptualised as being the result of an individual decision-making process. Likewise, a good deal of the conspiracy belief literature proposes vulnerability to conspiracy beliefs based on an individually-located drivers: a general tendency to believe these theories. Although, there have been calls for incorporating historical and societal context (Butter & Knight, 2015), and a recognition of the importance of group-based conspiracy beliefs in guiding those who design health interventions (Druckman et al., 2021), we have found no significant studies that focus on understanding the development of COVID-19 conspiratorial beliefs within EMGs.

## Method

This study examines COVID-19-related sensemaking in the UK Black community to develop theoretical insight into why conspiratorial explanations are more prevalent in this group. Semi-structured interviews were analysed based on a grounded theory methodology.

### Recruitment and participants

Twenty-eight participants were recruited as part of a larger project on COVID-19 behaviours in ethnic minority communities (see Vandrevala et al., 2022). To situate conspiracy beliefs within the Black community’s COVID-19 sensemaking we included the perspectives and experiences of members of the public, as well as those we refer to as community leaders or stakeholders, who were embedded within community groups, charities, faith organisations, and other organisations with high levels of attendance by the Black community. Of the 28 participants, 19 self-identified as Black African (15 were general participants, 4 community leaders) and 9 self-identified as Black Caribbean (7 were general participants, 2 community leaders). Our participant group is geographically diverse and draws from a variety of backgrounds. However, the nature of the study required that we restrict participation to those with a sufficient English understanding and speaking ability to undertake the interview and who were available to be interviewed by video or audio call. Consequently, our participant group was, on average, more educated than the general population. Therefore, we may have missed the representations of recent immigrants and those less technologically connected. However, for the purposes of theoretical development surrounding conspiratorial beliefs, our sample is an appropriate starting place. See author for detailed demographic characteristics.

After approval from the authors’ institutional ethics committee, initial contact was made with key individuals from a range of organisations, who then invited their members on our behalf. We encouraged prospective participants to contact us directly to arrange an interview, upon which we secured informed written consent. The researchers who ran the interviews and analysed the data were integrally involved in each stage (see Vandrevala et al., 2022 for further details on recruitment).

### Procedure and materials

We positioned conspiratorial beliefs as part of an individual’s system of sensemaking (Weick, 1995), with this sensemaking being subjective and socially constructed between the individual and society. The interview data is therefore seen to reflect an experiential and contextualised perspective.

The semi-structured interview schedule included questions that centred on the effects the pandemic had on participants’ lives, on their community, and on their perspectives on COVID-protective behaviours, such as social distancing and engagement with testing, isolating and vaccines (towards the later stages of data collection). Interviews also explored the perception of COVID-19 information particularly around health guidelines and government messaging. Semi-structured interviews allowed the researchers to be theoretically sensitive during the interview (following interesting leads) and prompts were used to develop and follow the ways in which participants made sense of the pandemic.

The interviews were conducted between August and December 2020 by the author LA, a member of the UK Black community; promoting comfort and confidence for participants. Interviews ranged from 45-80 minutes and were conducted via Zoom/MS Teams (n=26) and telephone (n=2). Interviews were recorded and transcribed. Participants received a £25 amazon voucher. The period from August to December 2020 covers a period in which the UK appeared to be in remission (end of August) to a nation-wide lockdown (November), followed by a temporary release in December. The data therefore covers a varying range of perceived threat from the virus, as well as a range of government-imposed restrictions.

### Data Analysis

The transcribed interviews were uploaded to MAXQDA 2020 for analysis. We selected grounded theory due to the relatively limited research regarding conspiracy in ethnic groups and the lack of a ‘grand’ theory to explain this phenomenon (Payne, 2021). Our objective was to identify a set of inter-related concepts that would explain why members of the UK Black community might hold COVID-19 related conspiratorial beliefs.

An initial familiarity with the literature surrounding health inequalities and COVID-19 was undertaken before analysis. The analysis was conducted with distinct first-order and second-order phases. The first phase involved an interactive use of reflective notes (summaries of our individual memos and thoughts of what was being said) written down by three researchers and discussed in a series of group meetings. For example, we discussed how the individuals interacted with different information sources and others in their community to gain a sense of what was happening. This enabled us to collate our memos and reflective notes into shared interpretations and dynamic, intertwined open-codes.

Up to this point, the analytic work was largely a-theoretical and inductive. As we entered the second order phase, we purposefully identified codes relating to explanations of, and justifications for, actions and beliefs, to which broader categories were assigned. By drawing on literature and theory that explains how wider theoretical categories are formed (Gioia et

## Findings

The following findings section unpacks in more detail the model presented in Figure 1. The aim of this model is to theorise the development of conspiracy beliefs in the UK Black community.

**Figure 1:**
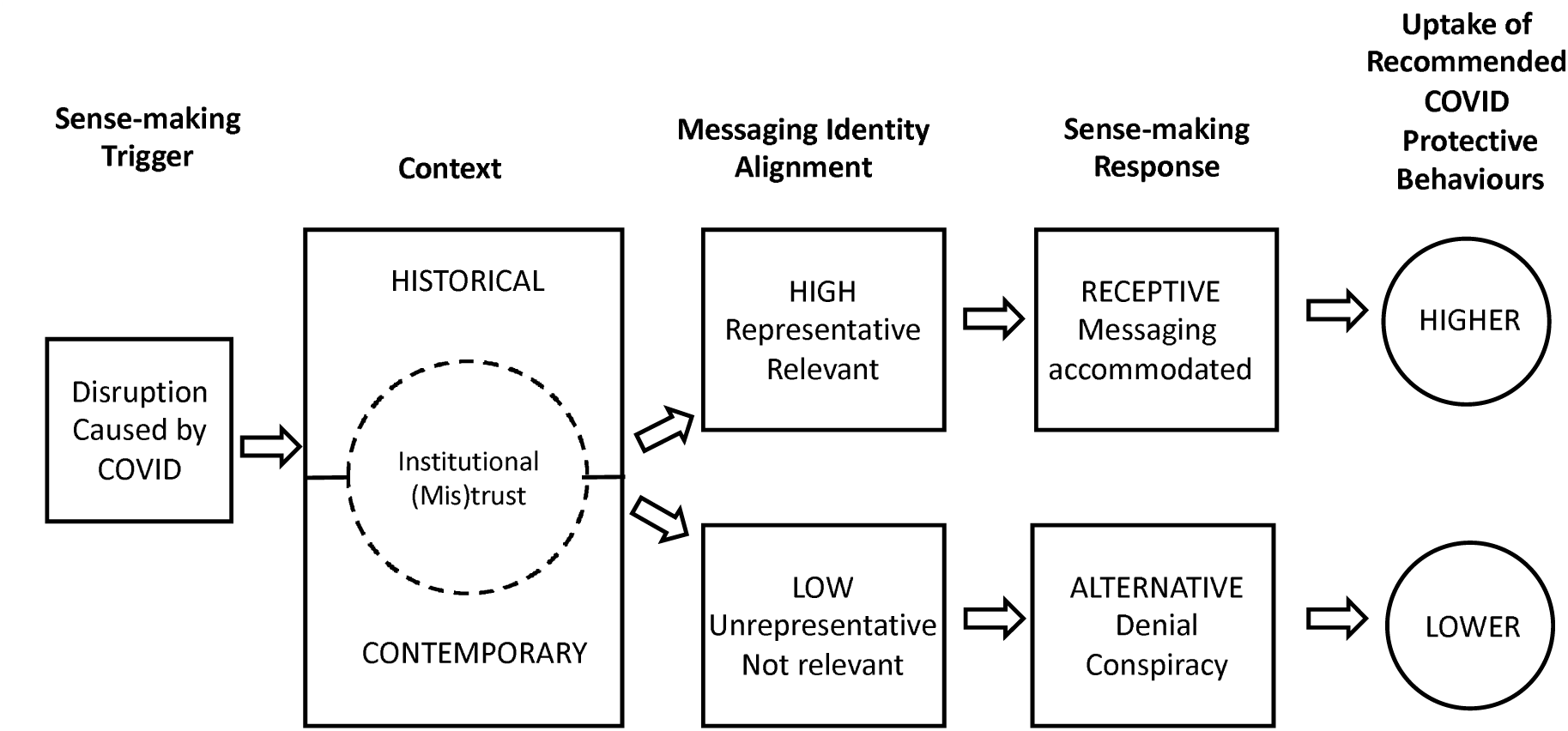
Theory to understand conspiratorial beliefs around COVID-19 in ethnic communities

Our participant group related narratives of sensemaking that were often reliant on conspiratorial beliefs which supported a lack of engagement with health protective Covid behaviours (such as engaging in testing or vaccine uptake). We found that this style of sensemaking was embedded in their lives within a context of institutional and societal mistrust, exacerbated by institutional messaging that was perceived as misaligned with their identity.

In the findings below we discuss the nature of the conspiratorial beliefs that our participants employed to make sense of the pandemic, and to explain decisions related to COVID protective behaviours. We highlight the integral role that messages during the pandemic played in promoting this type of sensemaking. Finally, we discuss the environment of mistrust, in which these alternative narratives rose and thrived. We found a particular distrust of the UK Government that could be traced to a wider environment of historical inequality.

### Sensemaking and conspiratorial beliefs

Sensemaking that employed conspiratorial beliefs generally followed one of two distinct themes; both of which supported non-engagement with protective health behaviours. The first theme held that the virus was either not real “*for me, it was just the myth*” (BA 5, female, 35-39 years) or was not as serious as it was being made out to be by the Government and media *“…a lot of the number have been fabricated in regard to the people dying”* (BC 6, female, 45-49 years). This particular conspiracy narrative was one of the more common themes abound within in the general public at this time (Freeman et al., 2020). In our respondents, these sensemaking beliefs related to the idea that the danger of the virus was being over-played and tied to participants’ everyday lived experiences. Conversely, this trust in everyday experience later provided a new validity to the seriousness of the pandemic, as the deadly consequences of the pandemic began to be personally experienced:

> *For me, it was just the myth…It was only when people that I knew of started to pass away. I spoke to my friend who caught it four weeks ago, and she spoke about her symptoms and how she’s still feeling a few weeks on, and it really made me think wow, okay, actually, this is real*. (BA 5, female, 35-39 years)

For some respondents a second, seemingly more sinister sensemaking theme included the belief that the virus was being wielded to intentionally harm particular segments of the population. This harm was not to particular individuals, or to society as a whole, but to certain groups:

> *I know other people also have other views about where this virus is coming from and whether it’s a way in which to control a particular population or it’s a way to erase a particular race from the surface of the earth*. (SBA 1, Male, 35-39 years)
>
> *I think it’s too coincidental that it is having such an impact on particular communities*. (SBC 2, Male, 55-59 years)

This ‘intentional harm’ belief goes beyond the typical conspiratorial assertions abound in the media at that time: that the Government and or ‘Bill Gates’ was attempting ‘population control’. This new belief was centred on targeting and harming very specific groups.

> *I have also heard that in our communities they think it’s a way of targeting the Black community, especially because of when it come out in the news that France was talking about having trial on Africa and it really did not give the communities much confidence in the purpose behind those kind of ideals* (BC 1, female, 30-34 years)

This narrative strongly suggests the existence of an environmental social uncertainty or threat for our respondents.

The connection between conspiratorial beliefs and engagement with protective behaviours was additionally evident when vaccines were discussed. In these accounts the Black community was seen as a target for Government misinformation and control:

> *So, in regard to, in the recent news and things, about vaccinations and things of that sort, they want to roll out trying out the vaccinations in our community before actually conducting correct trials in the correct way. Instead of saying, we’re going to try out in the African and Caribbean communities, which is not the way to go about things anyway, it’s supposed to be done with a diverse group of people from different communities. But obviously, there’s a little bit more to it…I think we’re disadvantaged that way. (BC 6, female, 40-49 years)*

For some this threat was further expanded as being within the healthcare system, in terms of purposeful neglect or mistreatment. The following exchange demonstrates shows how wider conspiratorial beliefs emerged within lived experience:

> Respondent: *A Black person goes to the hospital…Maybe we are just killing them. Maybe it’s that mentality, I don’t know…I believe that…Because there’s a lot of Black died during this Coronavirus. There’s a lot of Black African as a whole, a lot of Africans have died, including the Caribbeans… They don’t give them the same attention as they give the white people*.

### Misaligned messaging

Our respondents cited different forms of messaging as integral to the sensemaking narratives they held regarding COVID and protective behaviours. Respondents drew from evidence and made distinctions from messaging within their group of people and those from outside. Overall, the ‘outside’ messaging felt misaligned with their identity; positioning them as outsiders, and lacked the representation of messages that spoke to their lived experience. The unequal way in which the virus affected the Black community was clearly communicated by the Government and media during the first stages of the pandemic. This distinction acted to set the Black community apart from the majority population. This status as outsiders was exacerbated for some by the lack of black representation in the delivery of government health messages. One respondent communicates a common perception regarding representation:

> *I think the delivery of the messages, you have to involve somebody similar to them, isn’t it? Yes, to deliver that kind of message that look, I’m one of you. I’m like you, and this is how to do it, and they are likely to accept it, than purely white person. Let’s face facts, but people are likely to identify with somebody who looks like them, than somebody who doesn’t. (SBA 3, female, 55-59 years)*

While most respondents indicated that the message content was appropriate for everyone, others felt that messaging lacked recognition of difference, regarding the Black community.

> *So, the thing is a lot of people’s real experiences, it feels like all of these recommendations were made for people that live in that 2*.*4 kids or whatever. So, just man and woman, husband and wife and two children. And maybe they’ve got a mum and dad. But it’s not thinking about multiple families. (BC 5, Male, 40-44 years)*
>
> *Don’t link Asian and Black together, that whole BAME thing is ridiculous. It angers a lot of Black people. I’ll tell you that I will not read it, I will not look at it. So, we need to stop grouping Black and Asian people. A very strong opinion of mine. (BA 6, female, 25-29 years)*

The messaging was perceived by some as casting the Black community as outsiders, and did little to increase trust, against a background of uncertainty surrounding the pandemic with the environment of an ever-changing and sometimes contradictory set of rules:

> *I think again what is said about the message changing, that doesn’t help either. People think well, there should be a consistent message and if there is not a consistent message then it makes them not trust that source so much. (SBC 1, female, 60-64 years)*

This uncertainty and lack of trust in Government messaging was exacerbated by the way evidence of the impact of the virus was presented (in a strictly scientific and statistical manner):

> *So, obviously, the way they see things is based on evidence, so they can have that way of thinking. But the people where I’m coming from, from Southeast London, those large black communities that live in council estates, they don’t see science. In fact, they’ve not finished school. So, you can’t really bring evidence to them that’s not part of their worldview or not part of their way of living, actually. (BC 5, Male, 40-44 years)*

In sum, government messaging during the COVID-19 pandemic served to further erode the already tenuous trust within the Black community by serving to re-enforce previously held perceptions of marginalisation. This distrust and its environs are discussed in the next theme.

### Trust and inequality

Sensemaking that included conspiratorial beliefs was embedded in a wider narrative of distrust, particularly targeted at the Government, the healthcare system and to a lesser extent, science. In respondents’ narratives, the uncertainty of the current environment was contextualised by historical mistrust and additional contemporary uncertainty specific to the Black community.

### Mistrust in Government

Unlike previous US-based literature, in which science and the healthcare system were often significantly implicated in the distrust that breeds conspiracy in the general population (Harrington et al., 2021; Jamison et al., 2019) our data clearly implicates the UK Government. This may be due to the unusually high-profile role that the Government had in advising on health-protective behaviours during the pandemic, as compared to previous studies surrounding HPV and flu vaccines. In some exchanges, however, the distinction between the institutions of government, healthcare, and science, were difficult to distinguish.

Specific to the pandemic, there was a perceived misalignment between the high level of risk to the Black community, communicated in government messaging (discussed above) and the low level of action being taken to alleviate this.

> *I think it should be acknowledged as well that people from African backgrounds are more at risk of the disease. And what’s going on to try and address that? Because it brings more trust when you say the things upfront, when you come out and say, we know this is a problem and this is what we’re doing to address it, as opposed to not saying anything. I think most of it is just a trust thing, really, just to make sure that the people reading it trust the advice. I think that’s the main things, I would say. (BA 12, Male, 25-29 years)*

The gap between messaging and action is apparent, and how this gap makes space for alternative sensemaking to thrive. This gap by further grounded by a general distrust of politicians:

> *I think people are just always sceptical of politicians anyway. They just generally think that they’re corrupt. They’re liars. They don’t support us, and they don’t support people in the cause. They just ask for the votes and then we’re just scammed, basically. So, already with COVID coming in they were already fighting a lost cause. (*BA 5, female, 35-39 years)
>
> *The message doesn’t go across from these people. They don’t trust them, like I don’t. We came from a country that we don’t trust politicians because we think that they are not saying the right things. That’s why. (BA 7, female, 50-55 years)*

This last passage points to a historical mistrust based on experiences in the respondent’s native country. In this assertion is the implication that historical mistrust has not been alleviated by being in the UK. The above discussions regarding perceived otherness, and a lack of black representation point to both historical and contemporary perceptions of a lack of care by the UK Government, underpinned by recent negative experiences (such as the 2018 Windrush scandal (Gentleman, 2018) for Black Caribbean people):

> *At the moment, they seem to be a bit sceptical. Because unfortunately, I’ve noticed as well, there’s an item about the Windrush. Maybe because that was not being handled very well fact is now with the vaccine, a lot of black people are concerned that this is what I think is stumbling them. (BC 7, female, 55-59 years)*

### Wider historical and contemporary societal influences/inequalities

Awareness of historical racism was further highlighted by contemporary events such George Floyd’s 2020 murder in the US, and the Black Lives Matter movement (Booth, 2020), but for others, this perceived otherness was represented as being embedded in a lifetime of lived inequality.

> *How do I feel? I do feel that recent events have started to make me feel unwelcome here. Looking at social media, Black Lives Matter and all the protests and things like that. (*BA 5, female, 35-39 years)
>
> *To be honest with you, we are different and our hair is different, we talk different…That was in the 70s. We are in 2020 and the same nonsense is going on now and even worse type of thing. It’s not just about the Black Lives Matter movement that has happened now. This is last year. This is every day my kids go through it. I’ve got two boys stopped by the police, is this your car? Yes, it’s my car. What do you do and all those kind of things. I’ve lived through it… That is the kind of negativity that it’s given me all my life that I’ve been here. (BC2, female, 55-59 years)*

Against the background of a lack of care and representation, and perceived inequality, other additional and individual uncertainties were common, like recency of migration and lower socio-economic status. When combined, COVID-19 these factors appear to have been a catalyst, serving to increase the negative impact of already existing issues, and creating an environment in which conspiratorial sensemaking can thrive.

> *Well, there are so many inequalities on the COVID-19 issue. COVID-19 is like an eye-opener. COVID-19 opened our faces to a lot of things that we, BAME communities have been suffering from, like inequalities in work, inequalities in so many things, in hospitals, as in health-wise. So COVID-19 has really opened our eyes to so many things. (SBA 4, Male, 35-39 years)*
>
> *As a result of COVID-19 a lot of racial injustices have been brought to the surface, whether it be people working within the NHS or whether it being you just being a black person generally within* society. *And I feel that COVID-19 has highlighted those injustices that I’ve always known has exist. (BA 6, female, 25-29 years)*

Our theory (see Figure 2) explains how an environment of crisis (caused by the pandemic) combined with contemporary and historical mistrust, perceived injustice and inequality provided a context in which alternative conspiracy narratives to thrive. The nature of these conspiratorial beliefs made more sense than institutional sources (such as the UK Government), which further shapes their decision-making, leading to engagement or non-engagement with COVID protective behaviours.

## Discussion

The current study aims to develop a theory to explain the process underlying the development of conspiratorial beliefs around COVID-19, within Black African and Caribbean communities in the UK. By employing a sensemaking paradigm, our model widens the lens beyond individual cognitive determinants, common in health behaviour and conspiracy belief models. Our proposed model accounts for social context, in which health messages are received and the process by which these are compared to lived experience to determine their plausibility. Lack of representation, alongside group-based perceived injustice, created incongruence between institutional messaging and lived experience that fuelled conspiracy beliefs.

Our findings highlight that conspiracy beliefs are highly prevalent and embedded in the sensemaking of the UK Black community. Our respondent group employed a number of *group-based* COVID-19 conspiracy beliefs in describing sensemaking surrounding the pandemic, which extended beyond population conspiracy beliefs. Beliefs that powerful others aimed to cause intentional harm to particular groups in society (themselves) through the virus, or through virus-protective measures, were highly prevalent. These beliefs are explained by our model through their congruence with context; historical and contemporary injustice and institutional mistrust, against which new information related to the pandemic was assessed. Such group-based conspiracy beliefs were, for many, more in keeping with their understanding of the world than messages coming from institutions.

Individuals, especially those from collective societies, draw heavily on socially shared norms and understandings within their community. One of these norms is that ethnic minority groups have been historically exploited or neglected by medical authorities and this has led them to regard vaccination as a means of control rather than a public health measure for their good (Bish et al., 2011). This belief is additionally strengthened by perceived discrimination and distrust (Freimuth et al., 2017). The casting of BAME communities in the pandemic as high-risk, along with a new heightened awareness of otherness, as the result of the pandemic and other societal issues such as the Windrush scandal and the Black Lives Matter movement, is likely to increase perceptions of group□based deprivation and discrimination.

Our model speaks to the pivotal role of getting institutional messaging right. Institutional messaging from Government and public health agencies failed to find an anchor in terms of our respondent’s individual and group identity (Hendy et al., 2015; Vandrevala et al., 2022), and their lived experience. The extreme nature of the pandemic and the de-contextual nature of early government messaging (with an apparently excessive reliance of statistical science, and little context that related to black people’s day-to-day life) led to a feeling of exclusion and fear. Feeling of otherness was re-enforced by the reporting of higher risk to the “BAME” community with rules perceived to have been structured for more affluent, smaller households. The non-representation of black people in COVID-19 messaging served to re-enforce perceptions of otherness (Ahmed et al., 2019). This resulting alienation allowed alternate dialogues in the form of conspiracy theories to become an alternative way to make sense of the pandemic. Furthermore, the ebb and flow of the institutional messaging was perceived as confusing and lacked a consistent anchor for people from minoritised groups. so failed to create the needed connection with public health advice, further hindered by a residual context of perceived injustice and mistrust.

Previous literature highlights associations between mistrust in the institutions of government, healthcare, and science (Ðorđević et al., 2021; Harrington et al., 2021), with this mistrust rooted in perceived historical and everyday injustice (Hanson et al., 2021; van Prooijen et al., 2018). Our findings indicate that contemporary perceived injustice (including injustice outside of the UK) had significant influence. These findings mirror the theory posited by van Prooijen et al. (2018); that personal and group deprivation and experienced inequalities fuel conspiracy ideas. Our work goes somewhat further in providing detail of the interplay at work here; and how this subsequently impacted on sensemaking and the consequences of this for the uptake of recommended COVID protective behaviours.

## Limitations

These findings begin to illustrate the complex system that influences sensemaking in regard to protective health behaviours in the UK Black community. By their nature, these findings are specific to a particular minoritised group and moving forward, future research needs to test the transferability of our results. Although the model and the finding of the group-based conspiracy is likely to be applicable to other minoritised groups, the specific historical and contemporary influences that background those beliefs (and therefore point to how to address these) will vary across communities. Indeed, further analysis aimed specifically at understanding conspiratorial thinking may be able to prise influences specific to the Black African and Black Caribbean communities. Certainly, there are differences in influence on the conspiratorial environment, due to religious and cultural differences.

## Conclusion

The current study makes an important contribution to our understanding of how people make sense of the COVID-19 pandemic and why for certain groups in society their sensemaking is more likely to draw on conspiracy theory beliefs. Our findings suggest conspiracy theory beliefs prevented public health communication to hook into and anchor within individual, and group-based black identities. Given that conspiracy beliefs were not isolated or particular to a situation or a person, but widespread; stronger, groups-based interventions and strategies are needed to combat against communities being alienated from health messaging. In the past research, conspiracy theories were person and/or situation focused, but our study has highlighted that the global pandemic led to the rise of group-based conspiracy beliefs and provides useful explanations for why ethnic communities were so resistant to uptake of COVID testing and vaccinations.

Achieving sustained behaviour change relating to Covid protective behaviours (i.e., vaccination and testing) involves considering how specific groups understand new phenomena and the risks associated with them (i.e., their sensemaking and mental models of the phenomenon). Group-based conspiratorial beliefs are complex to tackle, and a group-oriented approach that considered contextual information relevant to EMGs, as this type of conspiracy stems widely from threat and uncertainty at a group rather than individual level.

Our work clearly indicates the importance of inclusion and representation in the source of the message given, with doubts more easily addressed though a grounded account from a trusted source, like testimony from community members and leaders. Awareness and acknowledging of the influence of past, endemic issues of discrimination, racism and injustice in health messaging and reflecting on ethnic minority communities lived experiences provides an opportunity to build trust (see Hendy et al., 2019). However, ensuring that community leaders, who themselves are members of the ingroup, are not the sources of misinformation and hesitancy is challenging. Community leaders have a vested interest in wanting the best for their community and hesitancy might come from them not knowing or having unanswered questions, and in the absence of this, spread information they think is right based on their previous, historic experiences (their own sensemaking). It’s important that information and messages are relayed to trusted sources in a timely manner (targeted information) to dilute the slew of the fake news and misinformation that can cause hesitancy.

## Data Availability

The data that support the findings of this study are available from the corresponding author upon reasonable request.

## Acknowledgements

The authors would like to sincerely thank our community organisations and participants for their contributions.

---

**Figure 1:**
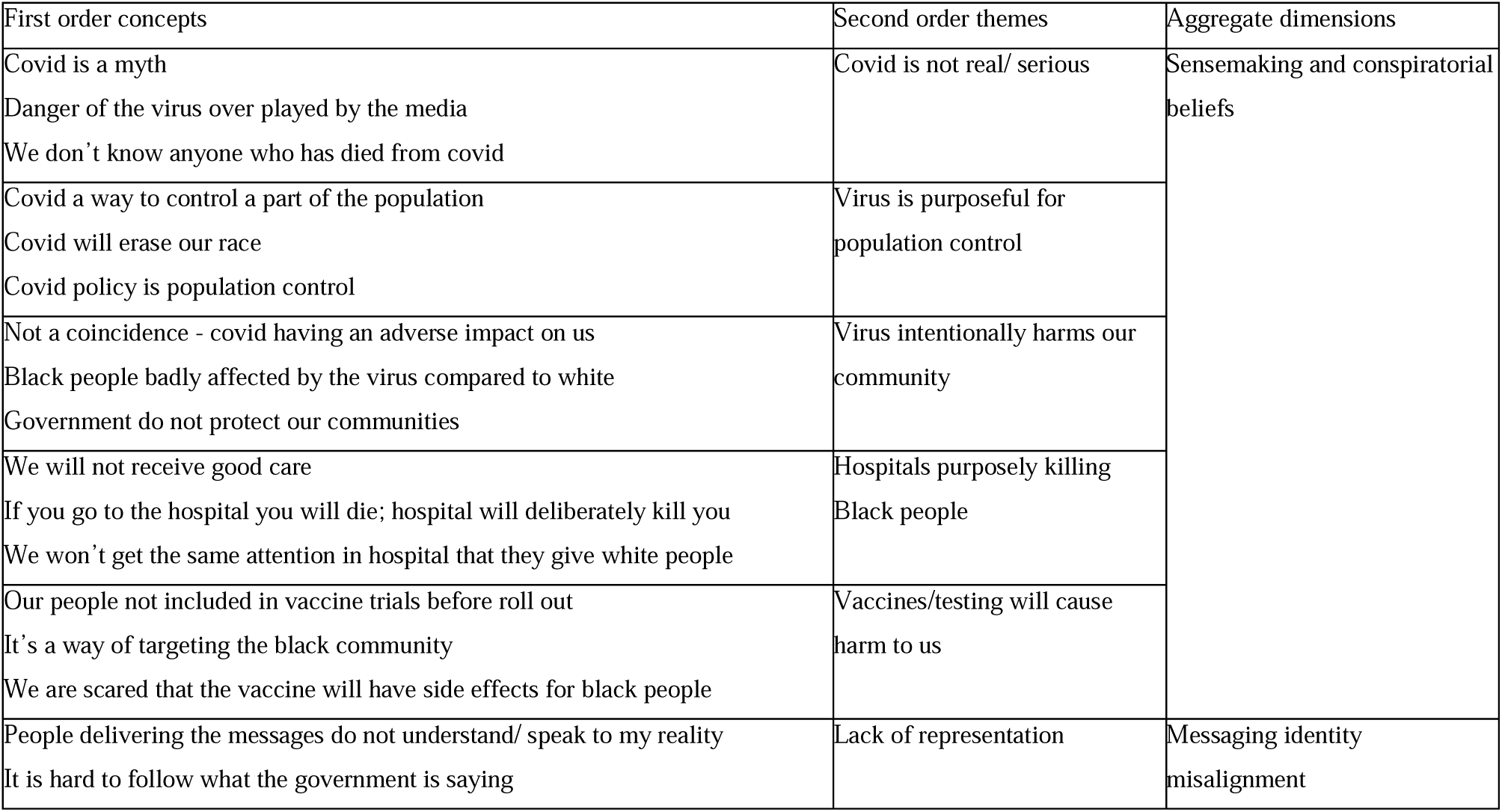
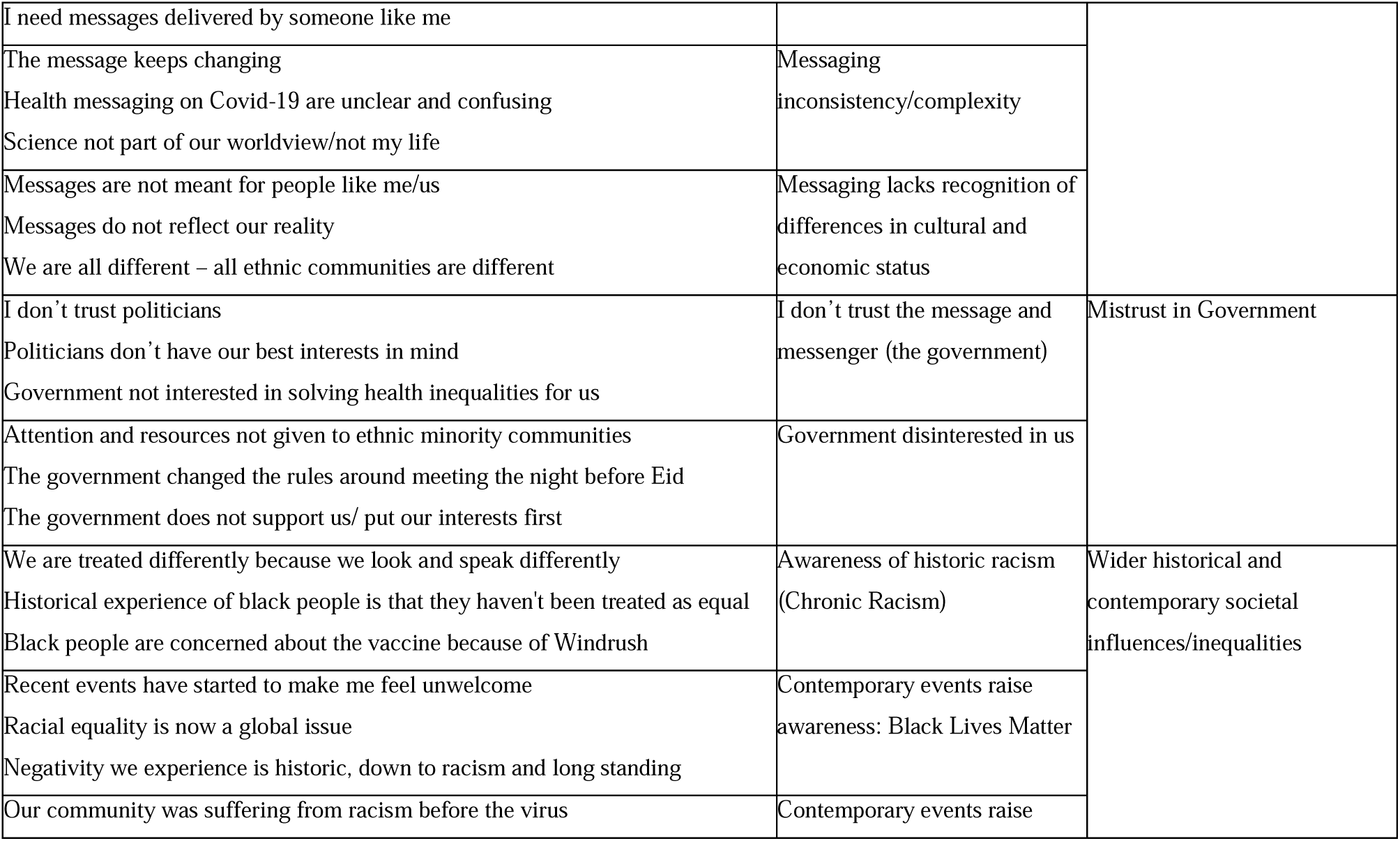
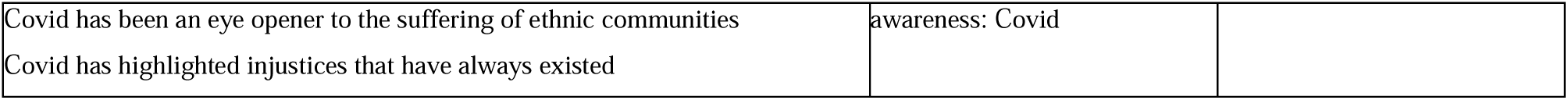
Data structure.

